# Autoencoder-based Feature Extraction and Classification for fMRI-based Deep Brain Stimulation Parameter Optimization for Parkinson’s Disease Treatment: Towards a Rapid Semi-automated Stimulation Optimization

**DOI:** 10.1101/2024.01.11.24301179

**Authors:** Afis Ajala, Jianwei Qiu, John Karigiannis, Brendan Santyr, Aaron Loh, Jürgen Germann, Desmond Yeo, Luca Marinelli, Thomas Foo, Alexandre Boutet, Radhika Madhavan, Andres Lozano

**Author notes:** Corresponding authors: A. Ajala. These authors contributed equally to this work (Afis Ajala, Jianwei Qiu).

## Abstract

Optimized deep brain stimulation (DBS) is fast becoming a therapy of choice for the treatment of Parkinson’s disease (PD). However, the post-operative optimization (patient clinical benefits are maximized and adverse effects are minimized) of the large number of possible DBS parameter settings (signal frequency, voltage, pulse width and contact locations) using the current empirical protocol requires numerous clinical visits, which substantially increases the time to reach optimal DBS stimulation, patient cost burden and ultimately limits the number of patients who can undergo DBS treatment. These issues became even more problematic with the recent introduction of electrode models with stimulation directionality thereby enabling more complex stimulation paradigms. These difficulties have necessitated the search for a biomarker-based optimization method that will streamline the DBS optimization process. Our recently published functional magnetic resonance imaging (fMRI) and machine learning-assisted DBS parameter optimization for PD treatment has provided a way to rapidly classify DBS parameters using parcel-based features that were extracted from DBS-fMRI response maps. However, the parcel-based method had limited accuracy as the parcels are based on subjective literature review. Here, we propose an unsupervised autoencoder (AE) based extraction of features from the DBS-fMRI responses to improve this accuracy. We demonstrate the usage of the extracted features in classification methods such as multilayer perceptron (MLP), random forest (RF), support vector machine (SVM), k-nearest neighbors (KNN) and LDA. We trained and tested these five classification algorithms using 122 fMRI response maps of 39 PD patients with a priori clinically optimized DBS parameters. Further, we investigated the robustness of the AE-based feature extraction method to changes in the activation patterns of the DBS-fMRI responses, which may be caused by difference in stimulation side and disease condition. Changes in the locations of activated and deactivated brain regions was simulated using a left-right horizontal flipping of the original left-sided (or nominal) DBS-fMRI response maps. The visualization of AE-based features extracted from the nominal and flipped DBS-fMRI response maps formed optimal and non-optimal clusters in a neuro-functionally meaningful manner, which indicate robustness of the AE-based feature extraction to subtle differences in the activated regions of DBS-fMRI response maps. The MLP, RF, SVM and LDA methods gave an overall DBS parameter classification accuracy of 96%, 94%, 92% and 93% respectively when trained using the AE-extracted features from the nominal DBS-fMRI maps. The AE-based MLP, RF, SVM and LDA accuracies were higher than the overall accuracy (81%) of our initial parcel-based LDA method. The performance of an AE-MLP model trained using the nominal DBS-fMRI maps did not change significantly when the model was tested on the flipped DBS-fMRI responses. We showed that the MLP method combined with AE-based feature extraction is best suited for fMRI-based DBS parameter optimization and represents another step towards a proposed digital tool for rapid semi-automated biomarker-based DBS optimization.

## 1. Introduction

Deep brain stimulation (DBS) is a neurosurgical procedure that involves the delivery of constant electric pulse using surgically implanted electrodes in specific target areas of the brain to suppress aberrant neural activities and/or modulate brain networks (1). DBS procedures are routinely adopted for the treatment of movement disorders such as Parkinson’s disease (PD), essential tremor and dystonia (2–4) and have additionally shown promising results for a range of psychiatric, cognitive, pain, and seizure disorders (2,5,6). The successful treatment of PD using sub-thalamic nucleus (STN) or globus pallidus internus (GPi) DBS hinges on precise surgical implantation and determining a patient-specific optimal combination of DBS parameters including signal frequency, voltage, pulse width and electrode contact location. DBS parameters are said to be optimized if they achieve maximal patient benefit while minimizing adverse effects (7). It is well known that sub-optimal DBS programming can lessen the treatment efficacy, increase patient side effects, and drain the implanted pulse generator battery more quickly than necessary (8).

Based on current standard-of-care empirical DBS programming, the search for an optimal combination of DBS parameters usually involves multiple time-consuming programming sessions that substantially increase the time to optimization (TTO) per patient (an average of 1 year), patient cost burden, patient fatigue and ultimately limits the number of patients who can undergo DBS treatment (9–11). Furthermore, the advent of newer DBS electrodes with an even greater number of directional contact locations renders the empirical optimization method increasingly difficult as the expanded DBS parameter space has made it intractable for clinicians to empirically program the electrode within a clinically acceptable timeframe. This increased complication and difficulty has hindered the adoption of the newer and more effective DBS electrodes by clinicians (12–16). In diseases such as dystonia, addiction and depression, clinically based programming is further complicated as there is a latency – potentially in the order of weeks – between stimulation adjustments and subsequent clinical effect (17). Due to such difficulties, an estimated 230,000 patients – a small number compared to the eligible patients for DBS – have undergone DBS therapy worldwide for a variety of neurological and non-neurological conditions (18).

The possibility of a biomarker-based DBS programming that can substantially reduce the TTO per patient during DBS therapy has been previously established by our research team (11,17). Optimal DBS parameter settings were shown to yield unique functional magnetic resonance imaging (fMRI) response maps in PD patients undergoing DBS therapy. Blood oxygenation level dependent (BOLD) DBS-fMRI response maps associated with optimal stimulation of the left STN showed significant deactivations in the ipsilateral motor cortex, contra-lateral cerebellum, orbito-lateral cortex, and significant activation in the ipsilateral thalamus. Subtherapeutic voltages triggered a decrease in the magnitude of the BOLD changes with a preserved topographic pattern. Supratherapeutic voltages yielded a relatively stronger BOLD response in the ipsilateral motor cortex and contra-lateral cerebellum and was also accompanied by increased BOLD signal in non-motor regions, such as the inferior frontal and occipital lobes.

Features that characterize the patient’s DBS parameters can be extracted from the DBS-fMRI response map and used to train a DBS parameter classification model via artificial intelligence (AI) methods such as multi-layer perceptron (MLP), random forest (RF), support vector machine (SVM), k-nearest neighbors (KNN), linear discriminant analysis (LDA) etc. In contrast with the current standard-of-care DBS optimization protocol, such AI models can facilitate the rapid classification of DBS parameter sets as either optimal or non-optimal.

The feature vector that is used to train the DBS parameter classification model may be composed of the entirety of voxels in the DBS-fMRI response maps. However, such full-voxel approach can substantially increase the length of the feature vector, model training time complexity and sensitivity to noise, without necessarily resulting in a superior classification accuracy rate (19). To avoid these difficulties, we had previously carried out our feature extraction using a parcel-based approach, where voxel intensities in predetermined brain regions of motor function relevance were extracted from normalized DBS-fMRI response maps (11). However, the accuracy and neurofunctional meaningfulness of this parcel-based feature extraction method may not be guaranteed as the parcels are based on subjective literature review and may not be able to take care of differences in DBS-fMRI responses that may occur due to differences in stimulation side and PD condition.

Here, in contrast to our previously implemented parcel-based feature extraction method, we propose an unsupervised autoencoder (AE) based feature extraction approach (free of ROI-based normalization) for obtaining feature vectors from the DBS-fMRI response maps of PD patients undergoing DBS therapy. The learned feature embeddings are subsequently used to train a DBS parameter classification model using the MLP, RF, SVM, KNN and LDA classification methods. We also compared the accuracy of the AE-based MLP, RF, SVM, KNN and LDA classification models to our previous parcel-based LDA model. Further, we investigate the robustness of the AE-based feature extraction method to changes in the activation patterns of the DBS-fMRI responses, which may be caused by difference in stimulation side and PD symptoms.

## 2. Methods

### 2.1. Experimental Data Description

In this work, we used our previously acquired BOLD fMRI data acquired on a 3T GE HDx MRI scanner (GE Healthcare, Wisconsin, USA) from 39 PD patients (n = 35 STN-DBS, n = 4 GPi-DBS, mean age = 62.4±7.1, 20 males, 19 females; study #NCT03153670) who had undergone left DBS treatment at Toronto Western Hospital. The optimal DBS parameters of all 39 patients was previously determined via the standard-of-care clinical optimization protocol (3,4). FMRI data was then acquired from all 39 PD patients with different DBS settings amounting to a total of 122 response maps. Data were acquired after protocols were approved by the institutional research ethics board at the University Health Network, #14-8255. All participants provided written informed consent prior to MRI scans, and a member of the clinical team was present to monitor patients during the MRI sessions. Patients were instructed to take their last dose of PD medication at least 24 hours before the study to avoid confounding responses. Other details of the patient demographics and DBS-fMRI data used in this work can be found in our previous publication (11). The BOLD fMRI design implemented in the study data was aimed at distinguishing patterns of brain activation at optimal and non-optimal DBS parameter settings. DBS-fMRI experimental data included 6.5-minute fMRI sessions using a 30 second DBS-ON/OFF cycling paradigm (Figure 1). All DBS-fMRI response maps correspond to different DBS parameter sets and were labeled as optimal or non-optimal by a movement disorder clinician based on optimal DBS parameter settings obtained via the standard-of-care clinical optimization protocol (3,4). Axial 3D anatomical T1-weighted images were also acquired for rigid-body registration. Other details of the DBS-fMRI and anatomical data acquisition parameters can be found in our previous publication (11).

**Figure 1.**
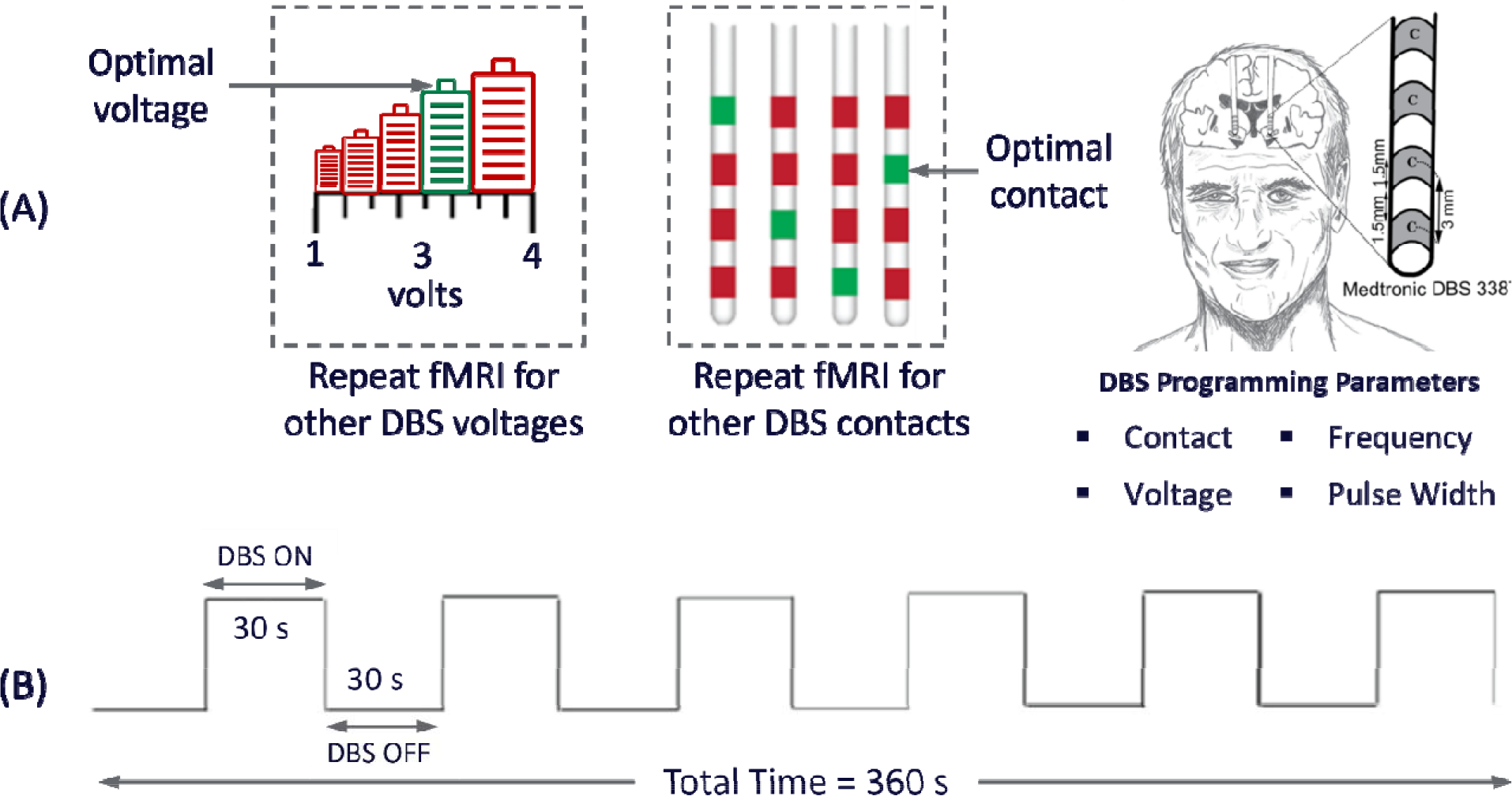
The acquired data consisted of fMRI data from left stimulated PD patients at optimal and non-optimal contacts or voltages (A). fMRI data were acquired using a 30 s DBS ON/OFF cycling paradigm for 360 s (B). The sketch of the human head was adapted from Boutet et al., 2021, as permitted under the Creative Commons Attribution 4.0 International License.

### 2.2. Single subject fMRI analysis

All fMRI data analyses were carried out using SPM12 (http://www.fil.ion.ucl.ac.uk). The acquired fMRI data were slice time corrected, motion corrected, rigidly registered to a T1-weighted image, non-linearly registered to a standard space Montreal Neurological Institute (MNI) brain, and spatially smoothed using a Gaussian kernel with a 6 mm full width at half maximum (Figure 2). To account for artifacts due to patient head motion during data acquisition, we used the Art toolbox (https://www.nitrc.org/projects/artifactdetect) (20) to detect and remove volumes with motion >2 mm. Overall, for any given patient, this resulted in the removal of a maximum of 6 volumes (3.3%) from the total number of acquired volumes. Motion regression was implemented in the fMRI design matrix using 6-degrees of motion (x, y, z, yaw, pitch and roll) before statistical parametric maps were extracted from the data. The 6-degrees of motion parameters were correlated with DBS ON/OFF block design to ascertain that the observed changes were related to DBS stimulation paradigm, and not related to patient head-motion. Statistical parametric maps (t-maps) were estimated from the preprocessed fMRI data using the designed 30-second DBS-ON/OFF paradigm. The data acquired for the first 30 seconds were discarded to establish a steady state. The hemodynamic response function was modelled using the canonical double gamma function, as it was found to be similar to BOLD fMRI response of DBS across brain regions and patients (20,21).

**Figure 2.**
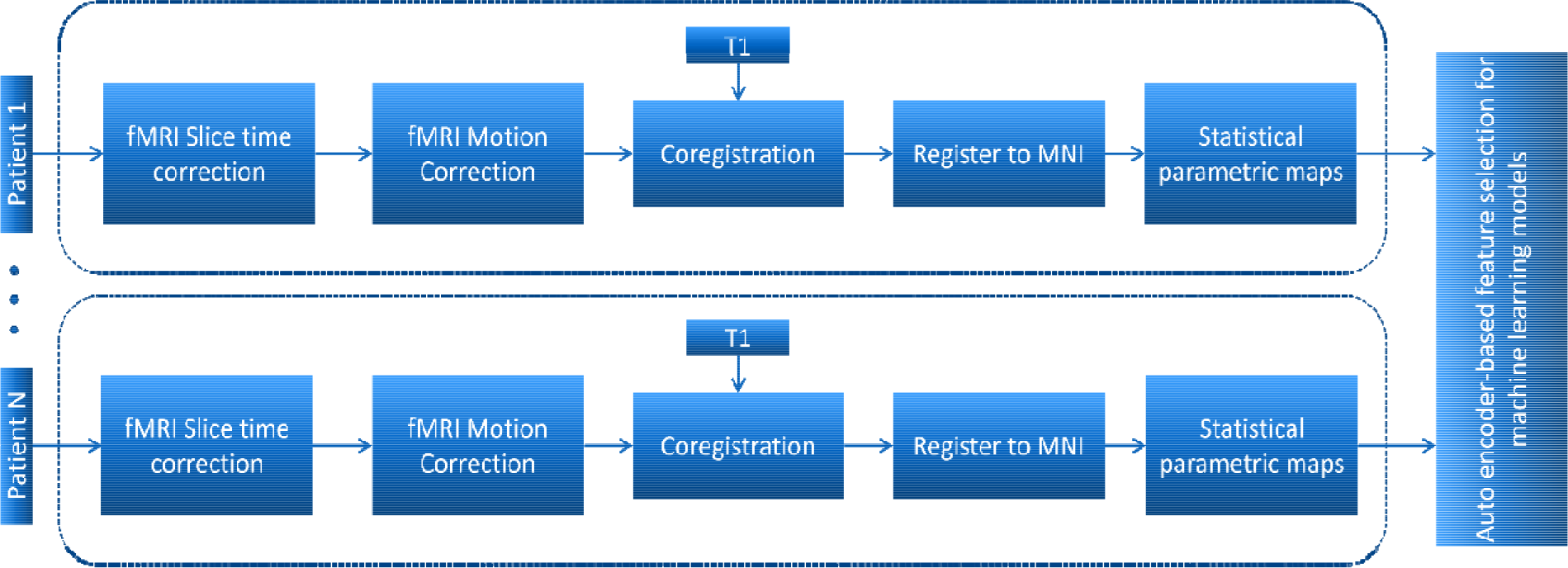
The acquired fMRI data from each patient are slice time corrected, motion corrected, coregistered to an anatomical (T1) image, registered to MNI space before carrying out a statistical parametric mapping analysis to obtain t-maps. The obtained t-maps are then passed to an auto-encoder network for feature learning.

### 2.3. Feature Selection Methods

#### 2.3.1. Parcel-based Feature Selection

In our previous parcel-based LDA implementation, features used for training the classification model were obtained from the average t-value in 16 ROIs of motor function relevance – that span the motor circuit, supplementary motor area, and cerebellum. These motor regions have been confirmed to be engaged during STN- and GPi-DBS (22,23). ROIs from other regions such as the operculum and visual cortex were included to account for speech issues and visual disturbances experienced with nonoptimal settings (24,25). Additionally, ROIs from other areas that could be related to common adverse effects (e.g., speech and gait disturbances) observed in PD-DBS patients at non-optimal contacts and voltages were included in the feature vector. The 16 functional-atlas-derived ROIs included (26): left and right thalamus, left and right pallidum, left and right sensorimotor, supplementary motor area, left and right anterior cerebellum motor, left and right posterior cerebellum motor, left and right operculum, primary visual, secondary visual and dorsal attention. This resulted in 32 features for each DBS-fMRI response map (16 positive and 16 negative mean t-values corresponding to neuro-activations and deactivations respectively) (11).

#### 2.3.2. Autoencoder-Based Feature Learning

The autoencoder is a neural network-based model that is used for unsupervised learning purpose to discover underlying correlations among data and represent data in a smaller dimension. The AE has a symmetrical network architecture, which is mainly designed to encode the input into a compressed and meaningful feature presentation (latent space), and then decode it back such that the reconstructed input (output) is similar or identical to the original input. A typical AE network is composed of two sub-networks popularly called encoder and decoder networks. The encoder is a compression function E that uses a kernel weight, activation function and bias (w, δ and b respectively) to map the input data x to a lower dimensional latent space z, which represents the latent space or bottleneck (27,28). The decoder is a recovery function D that maps the latent space z to the output x′ using kernel weight, activation function and bias (w′, δ′ and b′ respectively):

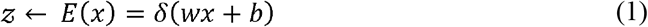

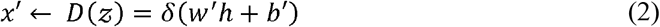

The goal of the AE network is to learn the mapping functions E and D:

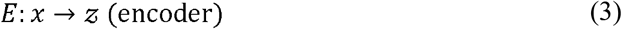

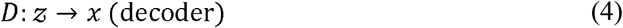

That satisfy:

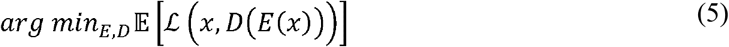

where 𝔼 is the expectation over the distribution of input x, and □ is the reconstruction loss function, which measures the difference between the original input and reconstructed input from the decoder (29). The goal is to minimize the difference between input and reconstruction through the defined loss function.

During the AE training process, the loss function is optimized using gradient back propagation. For simplicity and computational speed, L_2_ loss function such as mean square error (□ ^MSE^) is often chosen to compute the pixel wise difference (Equation 6).

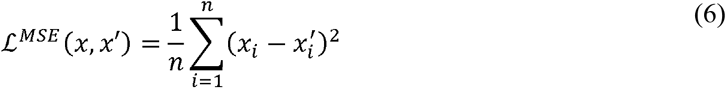

However, it is widely accepted that *L*_2_ loss does not correlate well with human perception of image quality as it does not capture the intricate characteristic of the human visual system (30). In this work, a perceptual loss function based on the structural similarity index measure (SSIM) was used for the AE reconstruction loss. SSIM captures the difference in luminance, contrast, and structural information instead of simply computing pixel wise difference. The SSIM is defined as (31):

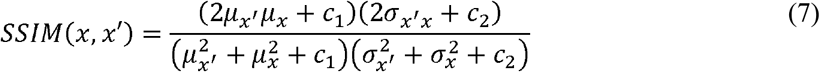

where μ′_x_, σ′_x_ represent the mean and covariance for x′ (similarly for x), σ_x_′ _x_ is the covariance of x′ and x. The values for c_1_, c_2_ stabilize the division with weak denominator. The SSIM-based loss function (ℒ ^SSIM^) is defined as:

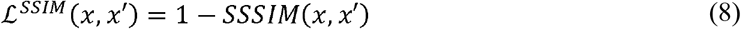

By optimizing this loss function, the network was able to reconstruct visually meaningful images that are perceptually similar to the original input images.

To extract associative features from the DBS-fMRI response maps without any prior information, we used the unsupervised convolutional AE network illustrated in Figure 3 to reduce the fMRI response maps from an initial dimension of 91 × 96 × 96 to a 256 × 3 × 3 latent vector, which has a length of 2304. All DBS-fMRI response maps were resized from the standard MNI dimension of 91 × 91 × 109 to a dimension of 91 × 96 × 96, which is suitable for our AE network. The resized response maps were also normalized between 0 to 1. The AE network produced reduced latent spaces that were used to form feature vectors for training the DBS classification models.

**Figure 3.**
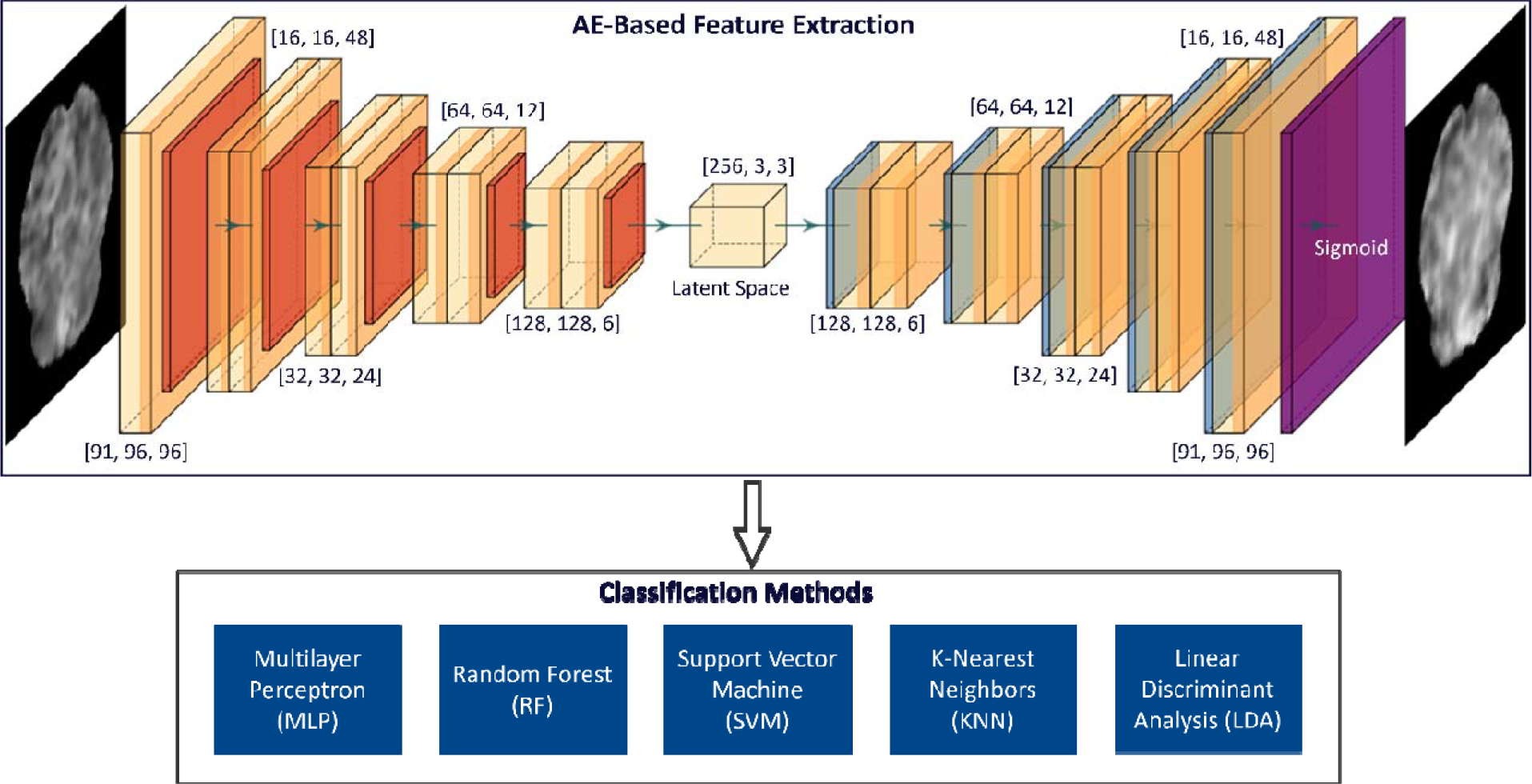
The network architecture for autoencoder (AE)-based feature extraction from fMRI response map feature learning (top). Number at each convolution block indicates the width, height, and depth of hidden feature map. The extracted AE-based features are then used to train AI models for the classification of the DBS-fMRI responses using the MLP, RF, SVM, KNN and LDA methods.

### 2.4. Artificial Intelligence Methods for DBS Parameters Settings Classification

In this study, we trained and tested five different AI models using the same feature vectors that were extracted (using the AE model) from the 122 DBS-fMRI response maps (32,33). The performance of the AE-based MLP, SVM, RF, KNN and LDA classification models were accessed to facilitate comparison with our previously published parcel-based LDA implementation (11). The KNN, RF, SVM and LDA distributions of the Python Scikit Learn package (34) were adapted in this work.

In brief, the MLP is a fully connected neural networks that functions as universal approximators and can be trained to approximate virtually any smooth, measurable function (35–37). Other details of the MLP has been explained in previous studies (38,39). Our implementation of the MLP method is composed of 8 blocks of hidden layers with a SoftMax activation function at the last layer to get the normalized classification probability for DBS parameter setting. Each neuron at any given hidden layer is fully connected to all neurons at the next hidden layer. Four dropout layers were added to the end of the first 4 blocks with feature dropout percentage of 25%, 15%, 15%, 15% respectively to prevent potential over-fitting. RF is an ensemble classifier that randomly builds multiple decision trees, that recursively partition data samples into two or more groups based on a specific splitting criterion such as Gini Index, Information Gain, and Gain Ratio (40,41). Other details of the RF method has been explained in previous studies (42–44). An SVM is a supervised AI method that defines a hyperplane to maximize the distance between any point in the training set and the defined hyperplane for accurate classification. The hyperplane is a decision boundary expressed in terms of a linear combination of functions parameterized by support vectors (45,46). More details about the SVM method have been previously published (47–49). The KNN is a supervised non-parametric classification method where unlabeled test data are labelled based on their similarity (using a distance metric) to samples in the training data (50,51). The LDA is a supervised dimension reduction and classification technique that finds an orientation matrix for the reduction of high dimensional feature vectors belonging to different classes to a lower dimensional feature space such that the projected feature vectors of a class on the lower dimensional space are well separated from the feature vectors of the other classes (52). We adopted the singular value decomposition solver for our current LDA implementation since the AE extracted features is quite large (2304 features) compared to 122 observations in our data set. A summary of the model parameters used for the tested AI classification methods are shown in Table 1.

**Table 1.** Summary of model parameters used during the training of the multilayer perceptron (MLP), random forest (RF), support vector machine (SVM) and k-nearest neighbors (KNN) classification methods. RBF: Radial Basis Function.

| Methods | Parameters | Values |
| --- | --- | --- |
| **MLP** | Number of layers | 8 |
|  | Optimizer | Adam |
|  | Learning rate | 0.001 |
|  | loss function | Cross entropy |
|  | Activation function | ReLU |
| **RF** | Number of estimators/trees | [2, 5] |
|  | Minimum sample split | [0.06, 0.20] |
|  | Maximum depth | None |
|  | Minimum sample leaf | 1 |
|  | Criterion | "gini" |
|  | Maximum feature | "Auto" |
| **SVM** | Kernel | Gaussian RBF |
|  | C (Regularization parameter) | 1 |
|  | Degree | 3 |
|  | Gamma | "scale" |
| **KNN** | Number of neighbors | [2, 5] |
|  | Weight functions | ["uniform", "distance"] |
|  | Algorithm | "Auto" |
|  | Leaf size | 100 |
| **LDA** | Solver | 'svd' |
|  | Absolute threshold | 0.0001 |
|  | Covariance estimator | None |

### 2.5. Classification Model Training and Testing

All model training and testing were implemented in Python 3.8 (Python Software Foundation, https://www.python.org/). The trained AE model was used to extract latent vectors from the 122 DBS-fMRI response maps, which were in turn used as features for training the MLP, RF, SVM, KNN and LDA classification models. Testing of the classification models was carried out within a 5-fold cross validation framework over 50 epochs. Eighty percent of the entire dataset was used for model training and the rest 20% was used for testing while maintaining stratified partitioning. The performance of all classification models was accessed using the receiver operating characteristics curves (ROC), accuracy, recall, precision and F1 score.

### 2.6. Latent Vector Visualization

To visualize the latent feature vectors learned by the proposed AE model from the 122 DBS-fMRI data, we applied the t-Distributed Stochastic Neighbor Embedding (t-SNE) method (53,54), which facilitates the visualization of high dimensional data by giving each data point a location in two or three-dimensional map.

### 2.7. Robustness of Autoencoder-based DBS Classification Model

Given that we used left-sided DBS-fMRI data for training our AE feature-extraction model, we investigated the robustness of the trained AE model to changes in the activation patterns of the input response maps. To mimic changes that can occur in DBS-fMRI responses because of differences in stimulation side and disease condition, the original left-sided (or nominal) response maps were passed through a left-right flip operation to displace the activated and deactivated regions horizontally yielding another set of 122 flipped response maps. Latent vectors were extracted from the flipped response maps using the same AE model that was trained on the nominal DBS-fMRI data. The latent feature vectors from the flipped response maps were separately used to train and test (also via a 5-fold cross validation) an MLP model for DBS parameter settings classification. The distribution of the latent vectors extracted from the nominal and flipped responses were compared using violin plots and cosine similarity index (CSI).

## 3. Results

The AE-based MLP, RF, SVM and LDA classifiers all showed a superior overall accuracy compared to our previously implemented parcel-based LDA method with a mean difference in overall accuracy of 13% (Figure 4). Only the KNN classifier showed a lower overall accuracy (80%) compared to the previously developed parcel-based LDA classifier with 81% overall accuracy.

**Figure 4.**
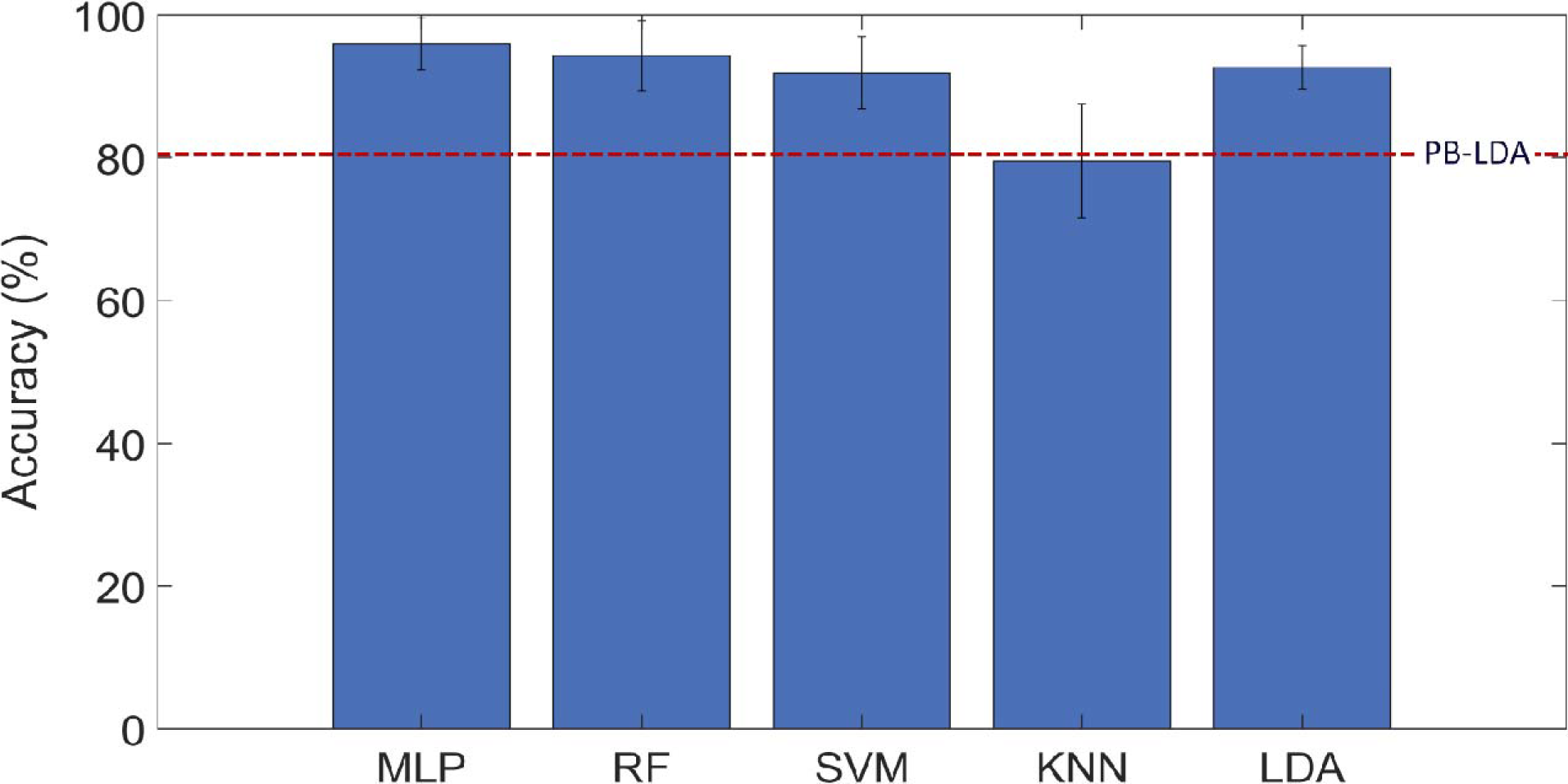
The mean accuracy of our previously obtained parcel-based linear discriminant analysis (PB-LDA) method is compared to those of the autoencoder-based multilayer perceptron (MLP), random forest (RF), support vector machine (SVM), k-nearest neighbors (KNN) and linear discriminant analysis (LDA) methods. Error bars represent the standard deviation across the 5-fold cross validation.

A comprehensive comparison of the mean and standard deviation (from 5-fold cross validation) of the accuracy, precision, recall and F1 score of the AE-based MLP, RF, SVM, KNN and LDA classification models (trained and tested on the nominal DBS-fMRI response maps) is summarized in Table 2. The MLP showed the highest accuracy, precision and F1 score (96%, 95%, and 93%, respectively) compared to the other classification models. The RF yielded the highest recall of all the five classification models. Following a similar trend, the ROC curves for optimal and non-optimal predictions were highest for the MLP classifier with an area under curve (AUC) of 0.98 (Figure 5). The AUC for the RF, SVM, KNN and LDA methods were respectively 0.96, 0.97, 0.79 and 0.97.

**Table 2.** Summary of accuracy, precision, recall and F1 score for the multilayer perceptron (MLP), random forest (RF), support vector machine (SVM), k-nearest neighbors (KNN) and linear discriminant analyses (LDA) classification methods. Parameter values represent mean ± standard deviation across the 5-fold cross validation.

| Parameters | MLP | RF | SVM | KNN | LDA |
| --- | --- | --- | --- | --- | --- |
| **Accuracy** | 0.96 ± 0.04 | 0.94 ± 0.05 | 0.92 ± 0.05 | 0.80 ± 0.08 | 0.93 ± 0.03 |
| **Precision** | 0.95 ± 0.07 | 0.87 ± 0.01 | 0.77 ± 0.12 | 0.67 ± 0.28 | 0.89 ± 0.09 |
| **Recall** | 0.92 ± 0.07 | 0.98 ± 0.04 | 0.97 ± 0.05 | 0.66 ± 0.18 | 0.87 ± 0.09 |
| **F1 Score** | 0.93 ± 0.06 | 0.91 ± 0.08 | 0.85 ± 0.06 | 0.63 ± 0.20 | 0.87 ± 0.07 |

**Figure 5.**
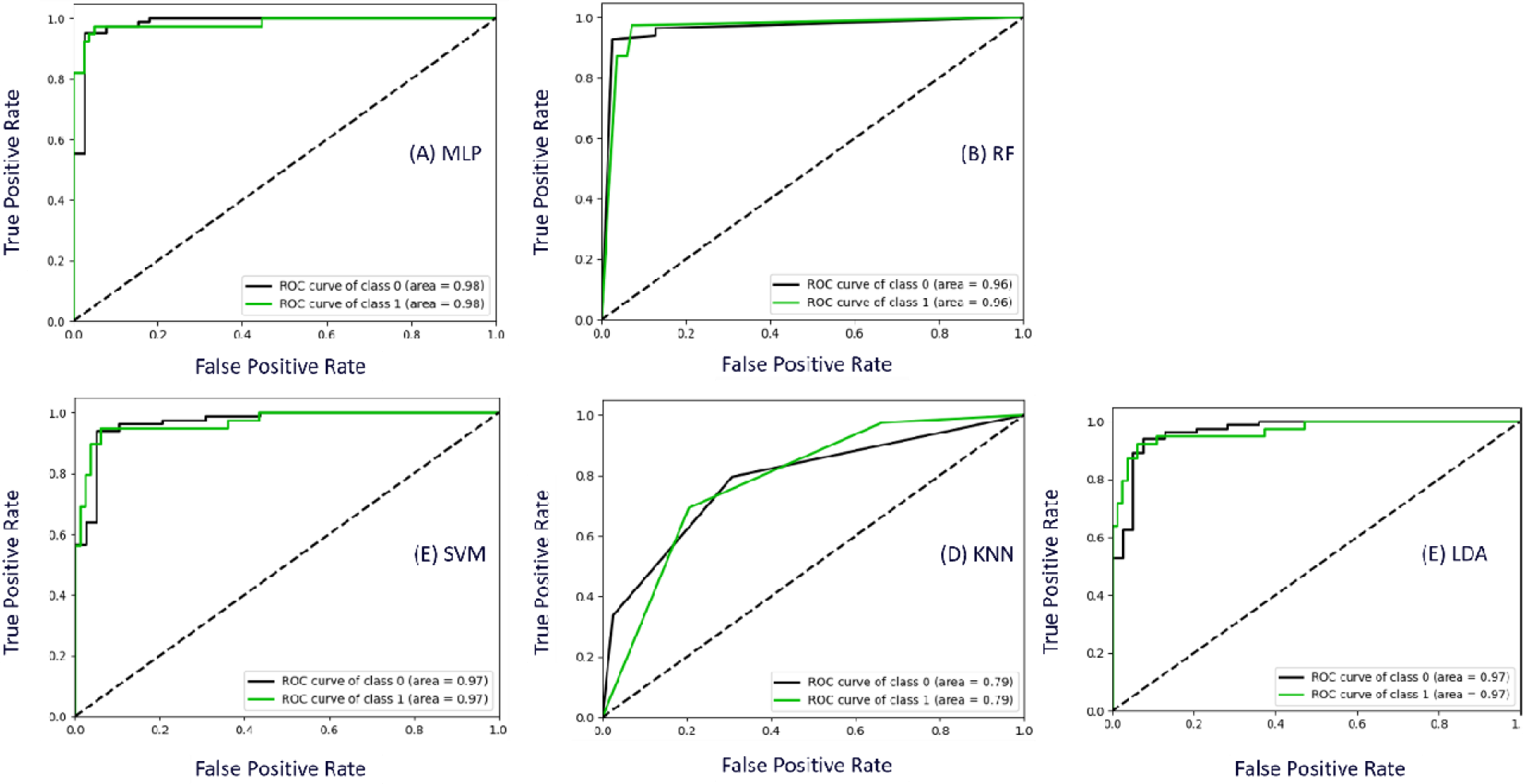
Results of ROC analysis for the multilayer perceptron (MLP) (A), random forest (RF) (B), support vector machine (SVM) (C), k-nearest neighbors (KNN) (D) and linear discriminant analysis (LDA) (E) methods with the AUC (area) displayed on the panels.

Representative nominal and flipped DBS-fMRI response maps, as well as the distribution of the AE-extracted features from optimal and non-optimal DBS-fMRI responses are shown in Figure 6A and 6B. Despite the difference in the topographic pattern of the nominal and flipped response maps, the distributions of their respective AE-extracted features were similar with CSI values as high as 0.783 and 0.683 for the optimal and non-optimal response maps respectively (Figure 6C and 6D). The t-SNE visualization of the AE-extracted features for the entire patient cohort (122 data points) were clustered in a neuro-functionally meaningful manner for the nominal and flipped DBS-fMRI response maps (Figure 7). Since the extracted features of the nominal and flipped responses are comparable, the accuracy of the AE-MLP classification model trained from both datasets was also similar with a difference of 4% (Figure 8).

**Figure 6.**
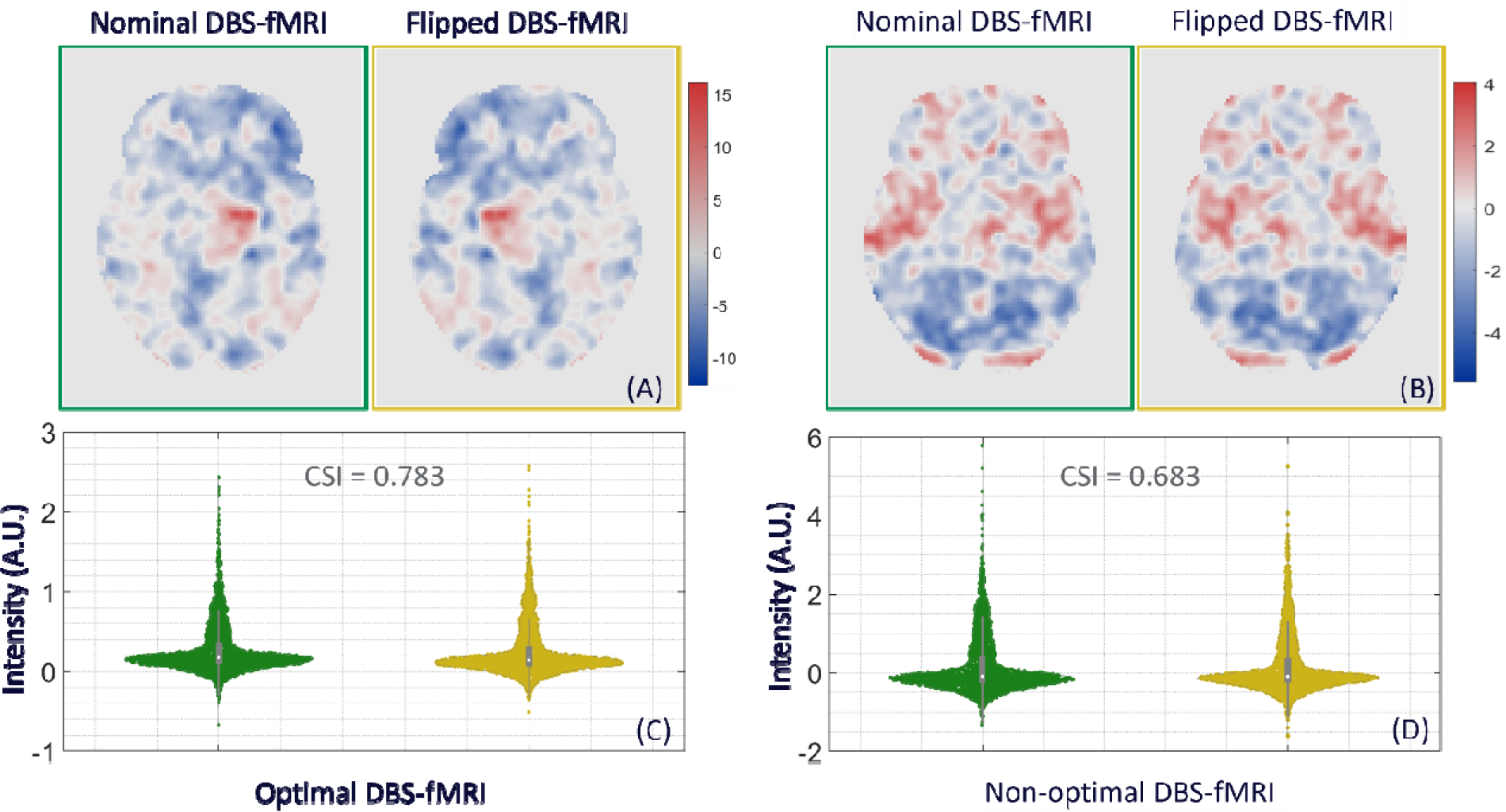
Axial view of representative nominal (green) and flipped (yellow) DBS-fMRI response maps at optimal (A) and non-optimal (B) stimulation parameters. The violin plots show the distribution of the features extracted by the autoencoder model (trained on left or nominal DBS-fMRI responses) from the nominal and flipped responses at optimal (C) and non-optimal (D) stimulation parameters. The cosine similarity index (CSI) of the features extracted from the nominal and flipped response maps are also shown between the violin plots.

**Figure 7.**
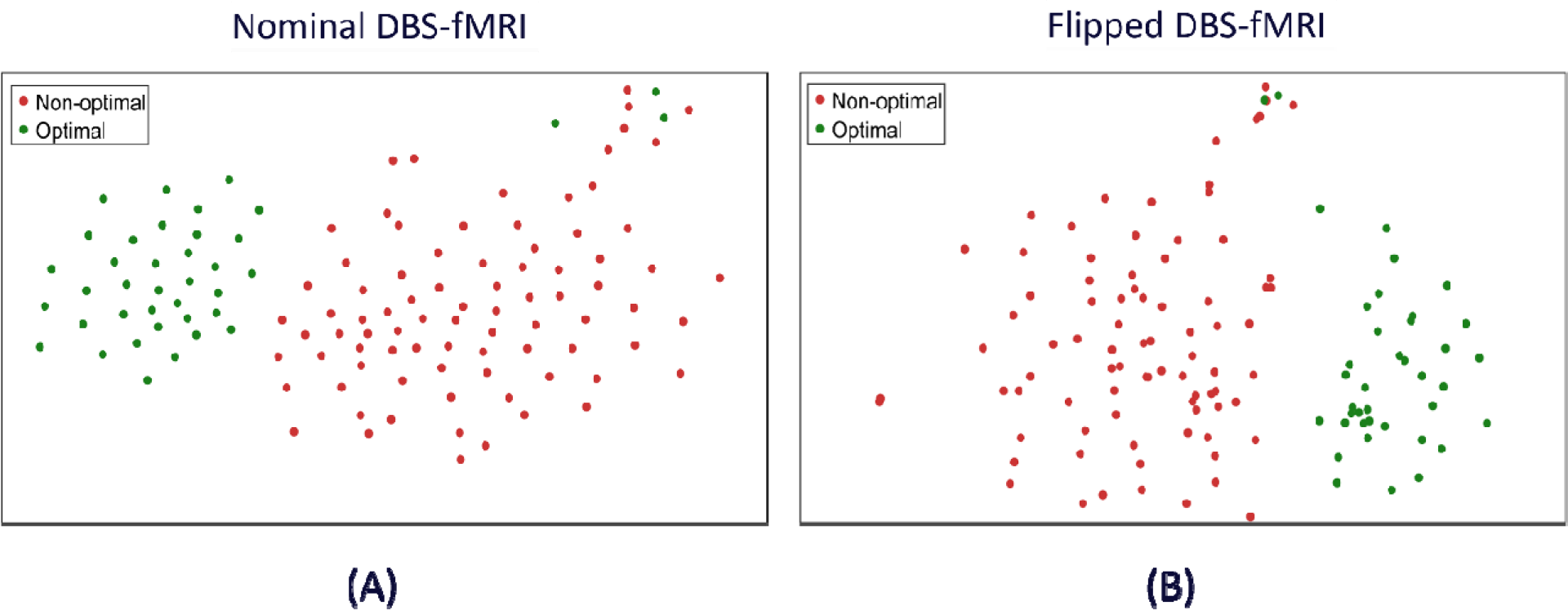
T-Distributed Stochastic Neighbor Embedding visualization of the latent vectors extracted from the DBS-fMRI response maps (of the entire patient cohort) using the autoencoder feature extraction model (trained on nominal or left DBS-fMRI responses only) indicate that the features obtained from the nominal (A) and flipped (B) DBS-fMRI response maps form clusters of optimal and non-optimal responses.

**Figure 8.**
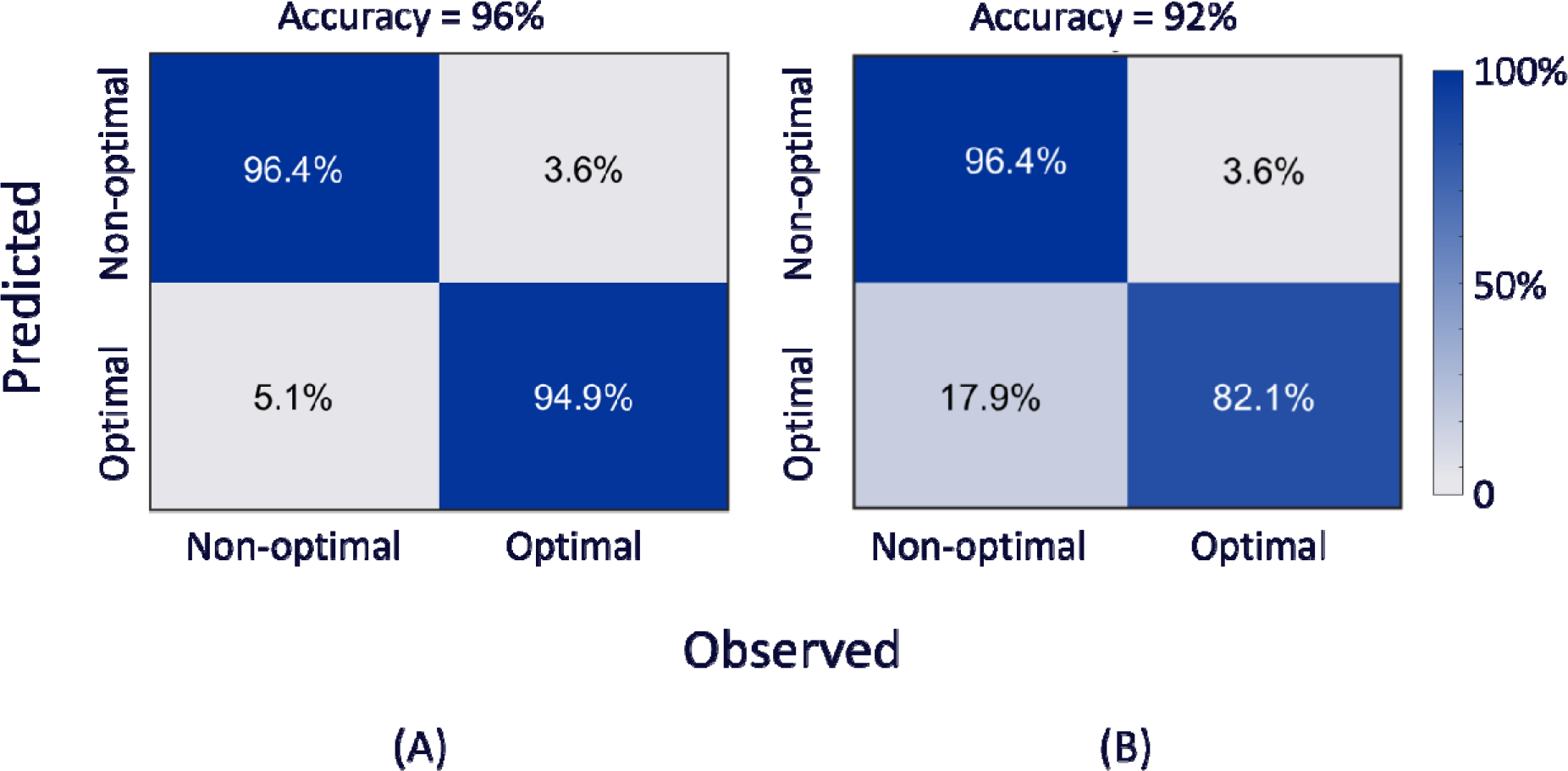
Confusion matrix of the autoencoder-based MLP classification model trained using features of the nominal (A) and flipped (B) DBS-fMRI response maps show comparable accuracies.

## 4. Discussions

In line with the recent search for a biomarker-based semi-automated rapid DBS optimization in PD patients, the AE neural network has been used as a feature extraction method for training MLP, RF, SVM, KNN and LDA classification models. In theory, the entire DBS-fMRI response map containing numerous features can be used for building the classification models, but doing so can quickly make calculations become laborious, less accurate and time consuming without a better understanding of the results (55).

The low false negatives and positives of the MLP model is a desired metric in the drive for rapid semi-automated DBS optimization and accounts for the high accuracy rate of the model. The AE-based KNN classification model gave the lowest accuracy, precision, recall and F1 score compared to the other AE-based classification methods, which may indicate that the KNN method is not suitable for classifying such high dimensional DBS-fMRI data. To further allude to the unsuitability of the KNN method, all other AE-based classifiers (MLP, RF, SVM and LDA) gave a superior accuracy rate compared to our previous implementation of the parcel-based LDA classification method except the KNN classifier (Figure 4). The use of the neuro-functionally meaningful features in the current AE-based LDA implementation yielded a significant increase in overall accuracy from 81% (for our previous parcel-based LDA implementation) to 93%, thereby highlighting the important role of an effective feature engineering method.

The inclusion of dropout layers in the MLP network prevented potential over-fitting and improved its performance in the adopted 5-fold cross-validation framework. Also, the use of SSIM loss function in the AE network proved to be more robust compared to using MSE loss function, as SSIM seeks to maximize the similarity between the input and reconstructed image (output) of the AE network.

The ROC curve analysis is also a good evaluative measure for determining the performance of various classification methods. We adopted the AUC to evaluate the performance of all five AE-based classifiers as it is widely used in medical research because of its meaningful explanation of the classification of various disease conditions (56). Among all four classification methods considered in our ROC analysis, the MLP method gave the optimal ROC with an AUC of 0.98, further showing the suitability of MLP for the classification of DBS parameter settings from fMRI data. The KNN performed the least with an AUC of 0.79.

The effectiveness of a single AE model (trained using only left-sided or nominal DBS-fMRI responses) is evident in the neuro-functional clustering of the extracted features as depicted in the t-SNE plots for the nominal and flipped response maps. This neuro-functionality of the extracted features broadly enhances the classification of the DBS parameter settings from their corresponding response maps. The variability in the optimal and non-optimal DBS-fMRI data is well captured in the eigen structures extracted from the response maps by the AE feature-extraction model (Figure 7).

Though our left-right flipping of the nominal DBS-fMRI response maps to generate the flipped responses does not exactly capture the real-world differences that will be imposed on a DBS-fMRI acquisition by difference in stimulation side or disease condition, these results may indicate that the AE-based autonomous feature extraction method is robust to differences in response maps that change the activated and deactivated regions horizontally. Our results also suggests that the left-right flipping operation may be used for data augmentation of single sided DBS-fMRI data, which is potentially useful for training more robust deep learning (DL) models for DBS parameter optimization.

A limitation of this work is the balance of the available data, which is composed of more non-optimal than optimal DBS-fMRI responses. However, the use of the AE-based unsupervised learning method may have reduced the effect of the data imbalance in the obtained results. The high F1 score of the models may suggest that the imbalance in our data is well tolerated by the AE-based MLP, RF, SVM and LDA classification methods, but not the KNN method (Table 2). Another drawback of this work is the limited number of available data set for model training and testing, which can be attributed to the relatively small number of PD patients that are able to gain access to DBS therapy (57–63). As more DBS-fMRI data become available, it may be possible to train a DL model that will predict optimal DBS parameters using features from a single fMRI acquisition as input, potentially reducing the TTO per patient further.

The present data was used to investigate fMRI brain changes associated with different DBS contact and voltage settings. The availability of fMRI data with DBS frequency and pulse width parameters will facilitate a more robust classification model for DBS parameter optimization. As previously demonstrated, fMRI is an effective biomarker of optimal DBS stimulation (11). Such biomarker-based programming tool could be leveraged to decrease the TTO per patient and number of clinic visits required before DBS patients’ optimal settings are identified. This is particularly important as the number of possible stimulation parameters increases with modern DBS electrodes that have been reported to be more effective with broader therapeutic window (64). The expanded parameter space of modern DBS electrodes has made it intractable for clinicians to perform DBS parameter optimization manually within a clinically acceptable timeframe, thereby hindering the adoption of these newer electrodes. Further, programming could theoretically be performed in the absence of specialized DBS physicians in non-expert centers.

Finally, the unsupervised AE-based DBS classification methods with accuracies as high as 96% represent another step towards a digital health care tool for semi-automated fMRI-based DBS programming. As such, we propose a rapid semi-automated DBS programming protocol that can substantially reduce the TTO per patient from an average value of 1 year (based on the current standard-of-care clinical optimization procedure) to 1 day during a single clinical visit (Figure 9). Such automatic DBS optimization systems could be transformative in diseases where there is a latency in clinical feedback (e.g., dystonia) (17), or where clinical responses are difficult to evaluate (e.g., depression) (65,66). Together, these benefits could dramatically increase the number of patients able to benefit from DBS therapy worldwide, while minimizing the time (and perhaps financial burden) needed for DBS optimization per patient.

**Figure 9.**
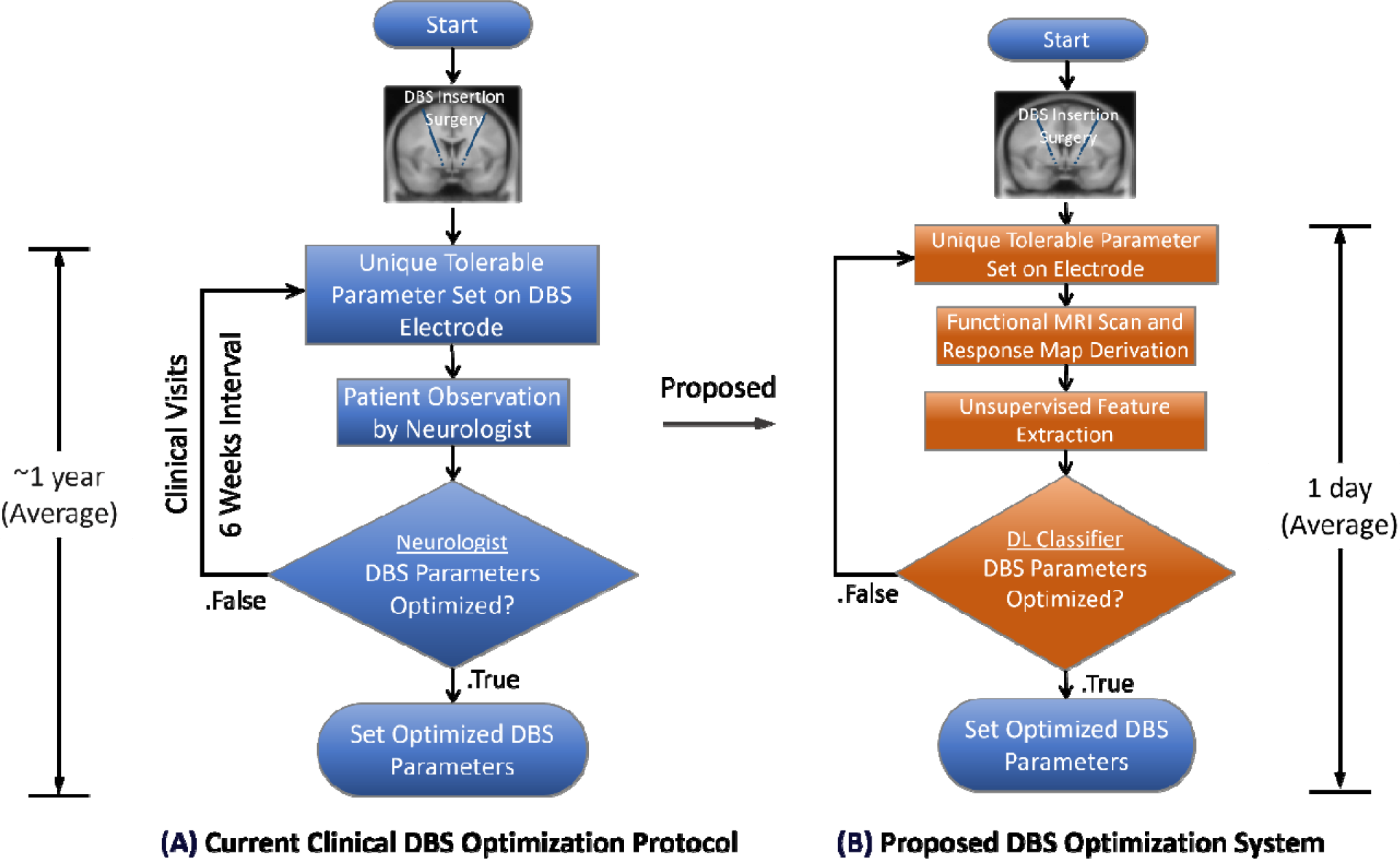
In the current empirical programming of DBS electrodes after the insertion surgery, the programming parameters (contact, voltage, frequency, and pulse width) are manually and sequentially adjusted until an optimal parameter combination is reached as determined by the neurologist (A). We propose a protocol that facilitates rapid optimization of DBS parameters using the fMRI and deep learning (DL) based feature extraction and classification models that were developed in this work (B). Such rapid semi-automated DBS programming protocol can substantially reduce the time to optimization per patient from an average value of 1 year (based on the current empirical optimization procedure) to 1 day during a single clinical visit.

## 5. Conclusion

Since fMRI has previously been shown to be a good biomarker of optimized DBS, we developed an autoencoder-based model for unsupervised feature extraction from DBS-fMRI response maps obtained from PD patients undergoing left-sided DBS therapy. We showed that the autoencoder-extracted features were neuro-functionally meaningful with robustness against subtle differences in the activated regions that may be caused by differences in disease condition and/or stimulation side during the acquisition of the training data. We then evaluated the performance of the extracted features in five AI classification methods. Among all five DBS parameter classification approaches tested, the AE-based MLP method gave the optimal accuracy, sensitivity, specificity, F1 score and AUC, which represents a much-desired improvement of our previously developed parcel-based LDA classification method for fMRI-based DBS parameter optimization.

## Data Availability

The datasets analyzed in this research are not publicly available due to data privacy regulations of patient data. Upon reasonable request, the study protocol and individual de-identified participants' raw fMRI data will be available to investigators from the corresponding author using private online cloud storage. Researchers wishing to validate or replicate this work using the same datasets would need to be approved by the research boards of University of Toronto, University Health Network and GE Global Research.

## Author Contributions

AA, JQ, JK and AL contributed to the conception, design, analysis, and interpretation of the data. AA and JQ drafted and revised the manuscript. AaL, JG, AB, and GE acquired clinical data and substantially reviewed the manuscript from a clinical point of view. RM, LM, DY and TF substantially revised the manuscript and gave critical analysis review points. AL supervised the study. All authors have read and approved the final version of the manuscript.

